# Tuberculostearic acid, a potential parameter for scoring system construction for Tuberculous meningitis diagnosis

**DOI:** 10.1101/2021.06.29.21259685

**Authors:** Tsz Hei Fong, Wangpan Shi, Siyi Li, Guanghui Liu, Chung Lam Ng, Haishan Jiang

## Abstract

This study aimed to validate the value of tuberculostearic acid (TBSA) whether it could implicate the existence of M. *tuberculosis* and assist for clinical diagnosis of Tuberculous Meningitis (TBM). Gas Chromatography/mass spectrometry was used to detect TBSA in the chemically pretreated cerebrospinal fluid of suspected TBM patients. In total, 140 patients were admitted for our study included 27 confirm TBM patients and 50 TBSA positive patients. Sensitivity of 0.7407 (CI 95%: 0.5372-0.8889) and specificity of 0.7345 (CI 95%: 0.6432-0.8132) were calculated. The Lancet consensus scoring system was also applied to evaluate the possibility of TBM in suspected patients, finding that TBSA positive patients showed a similar distributive grouping as the definite TBM patients. Our study implicates that the prospective use of TBSA is worth combining into a scoring system for characterizing the features of Mtb, showing a great potential of TBM diagnosis by TBSA in the future.

## Introduction

Tuberculosis meningitis (TBM) is the most dreaded type of extrapulmonary tuberculosis that developed in the central nervous system (CNS) in all kinds of tuberculosis (TB) diseases, although the average incidence remains low (5%∼15%) in all tuberculosis patients [1]. Hence, early diagnosis of TBM is essential to reduce the mortality and risk of poor prognosis for the patients [2].

However, one of the major issues concerning TBM is the low sensitivity of cerebrospinal fluid (CSF) microscopy and molecular assays [3], i.e. Ziehl-Neelsen staining, nucleic acid amplification test (NAAT) [4] and metagenomic next-generation sequencing [5], for the early diagnosis and follow-up of the disease, which complicates the early diagnosis and makes it a big challenge to prevent further neurologic damage [2]. As for culturing Mycobacterium *tuberculosis* (Mtb) from patient CSF, which is more sensitive than CSF smear microscopy, but requires liquid and solid media culture for at least 10 days and up to 8 weeks, respectively, as well as, ideally, a laboratory with biosafety level three [6]. Thus, all these direct methods for finding out Mtb, we deem as gold standard tests, have their strict limitations in early diagnosis for tuberculosis meningitis. On the other hand, the Lancet consensus scoring system has been well developed to improve TBM diagnostic accuracy, including various criteria such as clinical features, CSF findings, as well as neurological imaging [7]. However, this system also had limitations in bedside treatment decisions [8].

Tuberculostearic acid (TBSA) is a specific constituent of the cellular fatty acids of mycobacteria and could be considered serving as a biomarker in the Mtb detection in clinical specimens [9]. Traunmüller et al. showed that TBSA in plasma is valuable as a complementary diagnostic method in active tuberculosis because considerable amounts of TBSA are released into the blood circulation in pulmonary and extrapulmonary TB patients, providing additional information for suspected patients who present with tuberculosis related manifestation [10]. In TBM, some study showed that the gas chromatography/mass spectrometry (GC/MS) technique applied is highly sensitive for detecting CSF tuberculostearic acid in picogram (10 X-12g) amount [11, 12].

In our study, we aim to verify whether TBSA in CSF is a good sensitive marker for patients and a complementary diagnostic parameter for low sensitivity ‘gold standard’ in tuberculosis meningitis.

## Material and Methods

### Patients and controls

In this study, we have admitted 140 patients (age > 11 years) with symptoms and signs of Tuberculous meningitis including one or more of the following manifestations: headache, irritability, vomiting, fever, neck stiffness, convulsions, focal neurological deficits, altered consciousness, or lethargy. No patients were excluded and all of them were recruited and treated at Nanfang hospital and Zhujiang hospital in Guangzhou, China.

The patient’s CSF specimen was collected through the lumbar puncture and tested for TBSA with GC/MS. CSF direct culture (Löwenstein-Jensen Slope Medium), CSF smear microscopy (Ziehl-Neelsen staining), or NAAT (RT-qPCR using Mtb complex nucleic acid detection kit [Qiagen, Germany]) for Mtb were performed, which are known as the gold standards for TBM diagnosis, by Laboratory Medicine Center of Nanfang Hospital to distinguish the definite patients from patients with suspiciousness. CSF of each patient would be examined by at least one of the three tests. One positive result in one of the three assays found in the suspected patient was sufficient to determine as the ‘positive’ of TBM. All results of gold standards were provided by the Laboratory Medicine Center.

At the same time, we evaluated all patients by Lancet consensus scoring system [7] that requires the presence of symptoms or signs indicative of TBM plus additional 1) clinical, 2) CSF, 3) cerebral imaging criteria, and 4) evidence of TB elsewhere, with the determination of the most likely alternative diagnoses [8]. Four criteria contained a total of 20 parameters (Table 1) to score patients with different points of each parameter (maximum score: 20 pts) [7]. Suspected patients were classified by assigning scores into either ‘Definite (depend on gold standard results only)’, ‘Probable (>10 pts without imaging or >12 pts with imaging information)’, ‘Possible (6-9 pts without imaging or 6-11 pts with imaging)’ and ‘Not (<6 pts or with alternative diagnoses)’ TBM [7].

**Table 1.** Parameters in the Lancet consensus scoring system in Marais et al. (2010) (7).

| **Parameters in the Lancet consensus scoring system** | **Diagnostic score** |
| --- | --- |
| **Clinical criteria** | (Maximum category score=6) |
| Symptom duration > 5 days | 4 |
| Systemic symptoms suggestive of tuberculosis | 2 |
| History of recent | 2 |
| Focal neurological deficit (excluding cranial nerve palsies) | 1 |
| Cranial nerve palsy | 1 |
| Altered consciousness | 1 |
| **Cerebrospinal fluid criteria** | (Maximum category score=4) |
| Clear appearance | 1 |
| Cells: 10-500 /μl | 1 |
| Lymphocytic predominance (>50%) | 1 |
| Protein concentration greater than 1 g/L | 1 |
| CSF to plasma glucose ratio of less than 50% or an absolute CSF glucose concentration less than 2.2mmol/L | 1 |
| **Cerebral imaging criteria** | (Maximum category score=6) |
| Hydrocephalus | 1 |
| Basal meningeal enhancement | 2 |
| Tuberculoma | 2 |
| Infarct | 1 |
| Pre-contrast basal hyperdensity | 2 |
| **Evidence of tuberculosis elsewhere** | (Maximum category score=4) |
| Chest radiograph suggestive of active tuberculosis: signs of TB=2; miliary TB=4 | 2/4 |
| CT/MRI/ultrasound evidence for tuberculosis outside the CNS | 2 |
| AFB identified or Mtb cultured from another source (i.e. sputum, lymph node etc.) | 4 |
| Positive commercial Mtb NAAT from extra-neural specimen | 4 |

All patients were satisfied with the conditions of lumbar puncture and were informed consent for our research purpose and sample usage. The collected samples were kept in the CSF sample library of Department of Neurology in Nanfang Hospital. The study was approved by the Medical Ethics Committee of Nanfang Hospital (No. of approval letter: NFEC-2018-087) and supported by funding from the Science and Technology Planning Project of Guangdong Province, China (Grant No. 2017B020247006) as well as the Natural Science Foundation of Guangdong Province, China (Grant No. 2016A030313579).

### Chemical pretreatment and chromatographic conditions

CSF specimens were collected by lumbar puncture under aseptic conditions and stored in sterile test tubes. All CSF specimens were centrifuged (Centrifuge: Sorvall ST 16R, Thermo Fisher Scientific, USA) at 1006.2xg for 10 minutes at 4°C and 500μl of the supernatant was dispensed into a sterile cryotube, rapidly condensed in liquid nitrogen, and stored at −80°C. However, only 100μl of CSF from collected specimens were used for TBSA detection. 100μl of CSF sample was mixed with 300μl of methanol and 50μl of internal standard (0.3g/mL Nonadecanoic-d 37 acid) was added. After 15 seconds vortexing, 15 minutes of centrifugation at 25155xg was required. 370μl of the supernatant was transferred to a new EP tube and dried by blowing nitrogen at room temperature. Then 50μl of 20 mg/mL methoxyamine pyridine solution and Bis(trimethylsilyl)trifluoroacetamide (BSTFA) were respectively added to the blown sample, vortexed for 15s, reacted under 80°C for 15 min and blew by N_2_ for dryness. 100μl of hexane was added for 25155xg centrifugation for 5 minutes; the supernatant was extracted for GC/MS analysis.

Agilent 7890A-5975C GC/MS was used for chromatographic analyses. The used chromatographic column was DB-5 (25m×200μm×0.33μm) with 1μl injection volume and a split ratio of 4:1. Programmatic temperature rising with an initial temperature of 70°C, 10°C was raised per minute till 300°C, steady for 4 minutes. All data were collected by SIM mode with a scanning range of 40-600 and a solvent delay of 6 minutes.

### Statistical analysis

Statistical analysis was carried out by using GraphPad PRISM Version6.0c. Chi-square test was used for sensitivity and specificity analysis, *p* < 0.05 was considered significant.

## Results

In total, all 140 patients were admitted to undergo tuberculostearic acid detection in their cerebrospinal fluid, only 27 (27/140, 19.3%) of them were determined as confirmed Tuberculous meningitis through ‘gold standard’ examination. Each patient received at least one of the three ‘gold standard’ tests, finding that 20, 4 & 5 patients were having positive results in CSF smear (number of tests performed out of 140 patients: 133/140), CSF culture (67/140) and NAAT (96/140), respectively (Table 2). In which two of the patients with CSF smear-positive also showed a positive result in either CSF culture or NAAT. Among these positive tests, the corresponding numbers of positive TBSA results were shown in the brackets, indicating the tests with true-positive outcomes (Table 2).

**Table 2.** Stratification of ‘Definite’ (CSF smear, CSF culture and NAAT positive) patients into the Lancet scoring classification of ‘Probable’, ‘Possible’ and ‘Not’.

| **Lancet**<br>**Scoring** | **Definite (Gold standard)** |  |  |
| --- | --- | --- | --- |
|  | **CSF Smear +/-**<br>**Number of tests**<br>**performed**<br>**(TBSA +)** | **CSF Culture +/-**<br>**Number of tests**<br>**performed**<br>**(TBSA +)** | **NAAT +/-**<br>**Number of tests**<br>**performed**<br>**(TBSA +)** |
| **Probable** | 8/35 (6) | 1/18 (1) | 2/25 (1) |
| **Possible** | 9/80 (7) | 3/42 (3) | 3/58 (2) |
| **Not** | 3/18 (2) | 0/7 (0) | 0/13 (0) |
| **Total** | 20/133 (15) | 4/67 (4) | 5/96 (3) |

TBSA detection (0.5 nmol/L TBSA in CSF was selected as the cut-off level for a positive result, which is the smallest concentration of TBSA that could be detected by our GC/MS under our pretreatment method) found that 50 (50/140, 35.7%) and 90 (90/140, 64.3%) patients had positive and negative results, respectively. Table 3 illustrates that the patients’ distribution in terms of two different laboratory examinations. Among TBSA positive patients, 20 of them were found as definite TBM, obtaining the sensitivity of TBSA detection of 0.7407 (CI 95%: 0.5372-0.8889). Specificity, positive predictive value (PPV) and negative predictive value (NPV) were found as 0.7345 (CI 95%: 0.6432-0.8132), 0.4000 (CI 95%: 0.2641-0.5482) and 0.9222 (CI 95%: 0.8463-0.9682), respectively. Chi-square test was conducted for analysis, giving a significant difference *p* < 0.0001 between the gold standard group and the TBSA group.

**Table 3.** Comparison of the Gold standard examination and TBSA detection for patients’ CSF.

| **TBSA** | **Gold Standard** |  | **Total** |
| --- | --- | --- | --- |
|  | + | - |  |
| + | 20 | 30 | **50** |
| - | 7 | 83 | **90** |
| **Total** | **27** | **113** | **140** |

Note: gold standard includes tests of direct culture of cerebrospinal fluid (CSF), CSF smear microscopy , or NAAT for *M. tuberculosis*

In terms of the Lancet scoring system, regardless of grouping patients to the ‘Definite’, all patients were classified into ‘Probable’ (N=35), ‘Possible’ (N=86) and ‘Not’ (N=19) TBM groups (Table 4). ‘Gold Standard’ as well as ‘TBSA detection’ was respectively stratified to indicate patients’ distribution in each group that both methods showed a similar pattern of positive rates among groups (Highest: ‘Probable’; middle: ‘Possible’; and lowest: ‘Not’). Although ‘Possible’ showed a moderated positive rate due to including the largest portion of patients, it comprised the most TBM confirmed patients as compare to other groups. Also, patients in the ‘Not’ group could still be found with the positive results of gold standard (3 patients) and TBSA detection (4 patients). TBSA showed higher positive rates in the stratified groups and overall results than gold standard, indicating a stronger detection capability than traditional laboratory diagnostic assays for implication of Mtb existence.

**Table 4.** Comparison of the stratification of ‘Definite’ and TBSA patients.

| **Lancet**<br>**Scoring** | **N** | **Gold Standard**<br>**(Definite)** |  | **Positive rate**<br>**(number of +**<br>**patients/N)** | **TBSA** |  | **Positive rate**<br>**(number of +**<br>**patients/N)** |
| --- | --- | --- | --- | --- | --- | --- | --- |
|  |  | **+** | **-** |  | **+** | **-** |  |
| **Probable** | 35 | 10 | 25 | 0.286 | 17 | 18 | 0.486 |
| **Possible** | 86 | 14 | 72 | 0.163 | 29 | 57 | 0.337 |
| **Not** | 19 | 3 | 16 | 0.158 | 4 | 15 | 0.211 |
| **Total** | 140 | 27 | 113 | 0.193 | 50 | 90 | 0.357 |

N refers to the numbers of total patients within the stratified groups

## Discussion

In our best knowledge, no updated publications could be found for studying the diagnosis of TBM through tuberculostearic acid detection for cerebrospinal fluid since 2004. However, the methodology of TBSA detection in sputum or other body fluids still keeps updating for better diagnosis of pulmonary or non-CNS extrapulmonary tuberculosis. Therefore, we deem that the importance of TBSA for tuberculosis diagnosis in the CNS should be paid attention in order to develop an effective and valuable diagnostic evaluation system with TBSA.

In this study, we examined 140 patients’ CSF to find out the sensitivity and specificity of TBSA for TBM diagnosis. Our results showed differences from the first reported paper that had a promising sensitivity and specificity [11]. Despite the efficacy and accuracy of TBSA detection is lower than previous studies, this assay is a powerful near-direct method to find out the evidence of the existence of Mtb because TBSA is a cell wall component of Mtb [12]. We observed that TBSA detection always showed a higher sensitivity than gold standard tests, and had a similar pattern of patient distribution based on the Lancet grouping when not considering the ‘Definite’ group, giving clues that TBSA positive patients might contain the characteristics of the TBM confirmed patients shown (Table 4). TBSA positive can indicate the existence of Mtb in suspected patients who still cannot find direct proof for TBM through CSF examinations. Thirty patients in our study were seen as false-positive (Table 3), but it is possible that all (any) of them may be proved as real TBM in the future owing to the gold standard’s low sensitivity.

The Lancet consensus scoring system published in 2010 is universally acknowledged at the bedside for clinical judgment for suspected patients, and is conducive to treatment and prognosis evaluation [7]. However, we noticed that this scoring system could not provide adequate accuracy for the classification of undetermined true TBM patients when definite results of TBM could not be provided due to low sensitivity of gold standard tests. According to table 4, we found that a lot of definite TBM patients were grouped into ‘Possible’ and ‘Not’ rather than ‘Probable’ TBM, implicating that the efficacy of the Lancet scoring system still insufficient for physicians to decide the optimal treatment for patients. On the other hand, TBSA detection was found to have the distribution similarity as the gold standard and to have a higher sensitivity. If the detection results of TBSA could be deemed as the reflection for confirmed diagnoses, believing that more patients who were gold standard negative would be earlier treated correctly due to TBSA positive. Thus, we recommended that TBSA should be included as one of the evaluation parameters in the scoring system in order to characterize the feature of and compensate for the low sensitivity gold standard tests. TBSA is believed to have the great potential to provide the function for more accurate and earlier treatment for suspected TBM patients if it could be considered and taken as a significant variable in an evaluation system.

One of the limitations in our study is that patients did not undergo all of the three Gold standard tests due to the different clinical decisions made by different physicians and consideration of time consumption (e.g. CSF culture may require at least 4 weeks). Also, the false-positive patients consisted of true Mtb TBSA positive and other bacterial or fungal species TBSA positive patients [13]. The bacterial and fungal examination should be conducted for differential determination to exclude the false-positive patients who co-infected with other microorganisms. Besides, TBSA was the only analyzed molecule in our chromatographic analysis. Metabolomics analyses in chromatography study are expected to be carried out for searching multiple biomarkers in CSF for TBM diagnosis in the future.

In summary, we have obtained the sensitivity and specificity of TBSA detection in CSF based on 140 definite or suspected TBS patients and revealed the similar pattern of patient classification according to TBSA detection to the gold standard examination, illustrating the clues to create an evaluation system including TBSA that might enhance the diagnostic accuracy of TBM. An accurate scoring system, including TBSA detection outcome, needs to be designed in future research work to further verify the diagnostic value of TBSA.

## Data Availability

The data that support the findings of this study are available on request from the corresponding author, Haishan Jiang. The data are not publicly available due to the information could compromise the privacy of research participants.

